# Predictors of disease duration and symptom course of outpatients with acute covid-19: a retrospective cohort study

**DOI:** 10.1101/2020.06.05.20123471

**Authors:** James B. O’Keefe, Elizabeth J. Tong, Ghazala A. Datoo O’Keefe, David C. Tong

## Abstract

**Objective:** Describe the disease course in a cohort of outpatients with covid-19 and evaluate factors predicting duration of symptoms

**Design:** Retrospective cohort study

**Setting:** Telemedicine clinic at a large medical system in Atlanta, Georgia

**Participants:** 273 patients with COVID-19. Exclusion criteria included: (1) intake more than 10 days after symptom onset, (2) hospitalization for covid-19, (3) symptoms at less than two visits.

**Main outcome measures:** Symptom duration in days

**Results:** Common symptoms at diagnosis are upper respiratory (64% cough, 53% loss of smell or taste, 50% sinus congestion, 22% sore throat), systemic (50% headache, 48% body aches, 36% chills, 22% dizziness, 18% fever). The most frequent remaining symptoms at 30 days were cough (7%), loss of smell or taste (5%), body aches (5%), nasal congestion (5%), shortness of breath with exertion (5%), and joint pain (5%). Day of symptom onset was earliest for upper respiratory symptoms (mean 1.26 days, 95% confidence interval 1.15 to 1.4), followed by systemic symptoms (1.54, 1.39 to 1.7), with later onset of lower respiratory (2.86, 2.54 to 3.22) and gastrointestinal symptoms (3.46, 3.07 to 3.89), when present. Cough had the longest duration when present with 12.2 days (10.9 to 13.6). Loss of smell or taste had the second longest duration with 11.0 days (9.9 to 12.2). Provider-Assessed Symptom Severity (PASS) is the best predictor of symptom duration (P <0.005 for multiple symptoms) and patients with “Moderate” PASS compared to “Mild” at their intake visit have higher rates of symptoms at 30 days, including cough (12%), nasal congestion (10%), joint pain (10%), body aches (9%), loss of taste or smell (7%), headache (7%), and shortness of breath with exertion (6%).

**Conclusions:** Covid-19 illness in outpatients follows a pattern of progression from systemic symptoms to lower respiratory symptoms and persistent symptoms are common across categories. Provider-assessed symptom severity is the best predictor of disease duration.

## INTRODUCTION

Coronavirus disease 2019 (covid-19) has brought large numbers of patients to medical attention within a span of months for care of a previously undescribed illness. Early reports on the presentation and natural history of covid-19 appropriately focused attention on the severe cases and critically ill.[1-5] Subsequent surveillance has demonstrated that the majority of patients have milder forms of illness [6] and it is recommended that they remain at home with medical supervision.[7,8] Although the duration of home isolation is defined based on symptoms,[7] understanding of the symptom course of outpatients with covid-19 is limited and most reports include presenting symptoms alone or cross-sectional follow-up information.[9-17] Longitudinal symptom data and predictors of individual symptom duration have not been described.

In March 2020, we established a virtual clinic for the care of patients in home isolation with covid-19: the “Virtual Outpatient Management Clinic” (VOMC), using available knowledge for assessment and treatment guidelines. All patients underwent telemedicine intake visits with a physician or advanced practice provider (APP), including assessment of specific covid-19 symptoms using a standardized clinical note. Patients were followed for symptom management with regular telephone calls by registered nurses (RNs) and APPs until improvement or hospitalization.

As it became clear in clinical practice that symptom duration varies substantially between patients, we undertook this study to determine the predictors of symptom course of our VOMC cohort. We hypothesized that risk factors for covid-19 complications severity (demographics, comorbidities, symptom severity) would predict symptom duration.

## METHODS

### Study setting

The study was a retrospective cohort study, conducted at Emory Healthcare, the largest academic health system in Georgia (serving the greater Atlanta metropolitan area), which includes more than 250 provider locations and 120 primary care locations. The VOMC comprised an intake team of 14 physicians and 3 APPs from two primary care clinics; and follow-up call teams included 19 redeployed registered nurses (RNs) and 20 APPs. All intake providers were trained in the use of the risk assessment tool in a one-hour webinar and conducted a median of 25 intake visits during the study period (range: 5-99), with the majority of intake visits conducted by physicians (83.6%).

### Study cohort

We included outpatient adults who completed their VOMC intake visit between 24 March 2020 and 26 May 2020 with initial symptom dates between 17 March and 20 May. Exclusion criteria were: (1) intake visit more than 10 days after symptom onset, (2) hospitalization for covid-19 at any time, (3) symptoms at less than two visits (i.e. to be included, we required a minimum of one symptom reported during at least 2 separate visits, including intake visit and follow-up calls). We chose the exclusion criteria a priori in order to improve the accuracy of early symptom reporting and completeness of follow-up.

Subsets from this cohort have been reported elsewhere in a small case series[18] and for hospitalization risk prediction,[19] but the current study is the first to analyze complete longitudinal symptom reporting for the cohort.

During the study period, outpatient covid-19 testing was conducted by medical providers using nasopharyngeal sampling for real-time reverse transcription–polymerase chain reaction (RT-PCR) detection of severe acute respiratory syndrome coronavirus 2 (SARS-CoV-2). Adult patients with positive RT-PCR results from the screening clinics or emergency departments were called by a result notification team to provide self-care advice and refer for enrollment in the VOMC. The details of care are outlined in Box 1. Overall symptom severity was assessed by the Provider-Assessed Symptom Severity (PASS) criteria in Box 1. It was based primarily on respiratory symptoms.

#### BOX 1

**Virtual Outpatient Management Clinic Care**

##### Outpatient covid-19 testing criteria (March-April 2020)

1. Either (a) fever, cough, or shortness of breath or (b) two symptoms from the following: sore throat, congestion, myalgias, fatigue, diarrhea, loss of smell.
2. Prioritize: (a) frontline healthcare workers, (b) students on-campus and health professions, (c) CDC employees, (d) patients with risk factors (age, comorbidity, immunosuppression, work in a communal setting).

##### VOMC enrollment criteria

1. Diagnosis of covid-19 by nasopharyngeal PCR, <u>and</u>
2. Requiring outpatient management of covid-19 symptoms

##### Intake telemedicine visit

1. Documentation template includes symptom history, symptom severity (patient-reported and provider-assessed), past medical history, physical examination and risk assessment.
2. Symptoms assessed: “systemic” (fever, chills, body aches, dizziness, confusion, headache, joint pain), “upper respiratory” (loss of smell or taste, sinus congestion, sore throat, cough), “lower respiratory” (chest tightness, shortness of breath with exertion, shortness of breath at rest, wheezing), “gastrointestinal” (abdominal pain, nausea, diarrhea), and rash. Note: symptoms assessed as a single list and not grouped into categories during assessment.
3. Provider gives advice for (1) symptom management, (2) home isolation guidance and (3) outpatient monitoring.

##### Provider-Assessed Symptom Severity definition

1. Mild
  a. Respiratory: Cough, sputum production
  b. Systemic: Fever, chills, malaise, myalgia, anorexia, diarrhea, vomiting, headache
2. Moderate
  a. Respiratory: Severe cough, dyspnea on exertion, wheezing or sensation of mid-chest tightness
  b. Systemic: N/A (Not provided in VOMC clinical guideline)
3. Severe
  a. Resting dyspnea, labored breathing, resting pulse oximetry ≤92%, pleuritic pain, hemoptysis
  b. Systemic: acute confusion, severe weakness, syncope, acute decline in functional status

##### Follow-up phone calls (March-June 2020)

1. Patients receive follow-up telephone calls on the following schedule:
  a. Low risk: every other day for a minimum of 7 days from symptom onset
  b. Intermediate risk: daily for a minimum of 14 days from symptom onset
  c. High risk: twice daily for a minimum of 21 days from symptom onset
2. All patients called until the intervals above and for a minimum 3 days after improvement in fevers (without antipyretics) and improvement in respiratory symptoms (whichever criteria was longer).
3. Patients with improving or worsening symptoms could change tier after enrollment at provider discretion.

### Data sources

Study data were obtained from two specific provider notes types deployed in March 2020 within the Emory Healthcare electronic health record (Cerner Corp., Kansas City, Missouri, United States): (1) VOMC provider intake assessment and (2) VOMC follow-up telephone call. The intake assessment note template included (1) documentation of specific covid-19 symptoms including onset and offset dates, (2) patient reported and provider-assessed symptom severity (PASS), and (3) documentation of specific medical conditions associated with risk of severe covid-19 (based on medical literature search in March 2020). The follow-up telephone call template included an identical symptom list with “yes/no” selection for documentation of the presence or absence of symptoms at follow-up.

If symptom onset date was not identified in VOMC notes, we conducted manual chart review of telephone records prior to VOMC enrollment. Additional demographic information including age, gender, and race (if recorded) was included from the electronic health record.

To ensure that symptoms were counted only once a day per patient, among patients receiving 2 calls per day, if a symptom was listed as present more than once for a particular day, it was counted only once. Among patients receiving calls every other day, if a symptom was present on both the preceding and subsequent day it was listed as present on the single non-call day in between for symptom duration.

### Main outcomes

The main outcome was symptom duration in days. The outcomes were the durations for each specific symptom. For symptoms not present on all dates (i.e. waxing and waning), we used the first and last documented dates of the symptom to document duration. The secondary outcome was the day of symptom onset. Symptoms were groups into systems: upper respiratory (cough, congestion, sore throat, loss of smell or taste), systemic (body aches, chills, dizziness, headache, joint pain), lower respiratory (shortness of breath with exertion, shortness of breath at rest, chest tightness) and gastrointestinal (nausea, abdominal pain, diarrhea). Clinical record extraction was conducted 21 June 2020 at which time all enrolled patients had at least 30 days of follow up based on symptom start date and all patients had received their final VOMC nurse call.

### Bias

Screening criteria are noted in Box 1. Healthcare employees were prioritized in the screening process and may be overrepresented in the cohort. Patient enrollment in VOMC was voluntary at the time of results notification, which may result in selection bias. Patients were scheduled for the minimum recommended follow-up calls at the time of intake (and could later extend care further if needed) but could disengage on request, which could lead to attrition bias.

### Predictors

Demographics, comorbidities, patient reported symptom severity and PASS were tested as predictors of disease duration.

### Statistical analysis

Duration of symptoms (in days) was found to have a positive skew so a natural logarithmic transformation was used. This was compared to a gamma loglink, negative binomial with loglink and a square root transformation of duration. The natural logarithmic transformation was chosen because it had the smallest Akaike Information Criterion (AIC) and Bayesian Information Criterion (BIC). Mean durations and 95% confidence interval were calculated based on the log transformation and then exponentiated back to obtain the result in days. One-way ANOVA was used to compare the mean log durations for each symptom between different groups for each predictor. Because multiple comparisons are being made we were looking for p values < 0.005 rather than 0.05. Different predictors significant to this level were to then be included in a multi-way ANOVA. The results were then exponentiated back from natural log of days to days for results.

Similarly, the day of symptom onset organized by systems was found to be positively skewed. In this case a natural logarithmic transformation was also used because it had a smaller AIC and BIC than the gamma log link and negative binomial with loglink of start day. Square root transformation of start day had a slightly better AIC (2108 vs 2161) and BIC (2132 vs 2185) than the logarithmic transformation, but we used the logarithmic transformation to maintain consistency with symptom duration (which was transformed with a natural logarithm). ANOVA compared the day of symptom onset between the different systems (systemic, upper respiratory, lower respiratory and gastrointestinal). After analysis the natural exponent was taken of the results to present the result in day of symptom onset.

### Patient and public involvement

Patients and the public were not involved in the design and conduct of the study, outcomes, recruitment, or planned dissemination.

## RESULTS

551 intake visits were completed in VOMC between 24 March 2020 and 26 May 2020. We included 273 patients in the study after excluding: 123 patients with intake visit more than 10 days after symptom onset, 62 patients hospitalized at any time for covid-19, 57 patients with symptoms on less than two visits, 26 patients that did not receive follow up calls, 7 patients without documented positive RT-PCR test for SARS-CoV-2, and 3 patients with blank or uninterpretable symptom entries.

### Characteristics of the study population

Table 1 describes demographics, comorbidities, and symptoms at intake visit for the cohort. Since subsequent analysis showed only symptom severity as significantly predicting disease duration, we also describe our cohort by PASS. With all patients hospitalized for covid-19 excluded, we only had four patients with PASS “severe” in our study. All four were black women, and their only comorbidities were asthma, hypertension, and obesity. There was no statistically significant difference in age by PASS. Patients in our study had mean age 45.7 years, 69% women, and 47% black. The PASS groups differed significantly (p<0.05) for comorbidities asthma and immunosuppression.

**Table 1.**
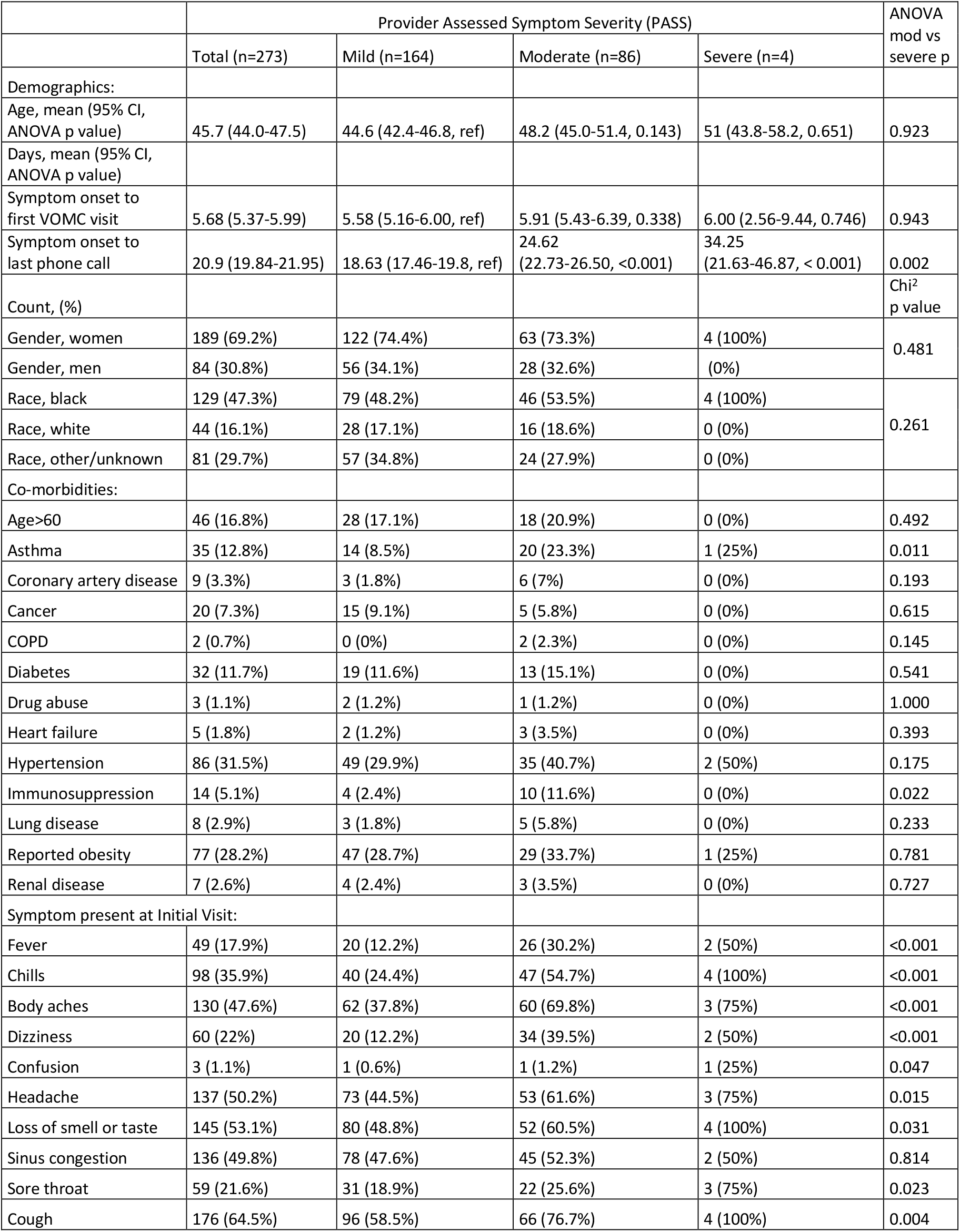
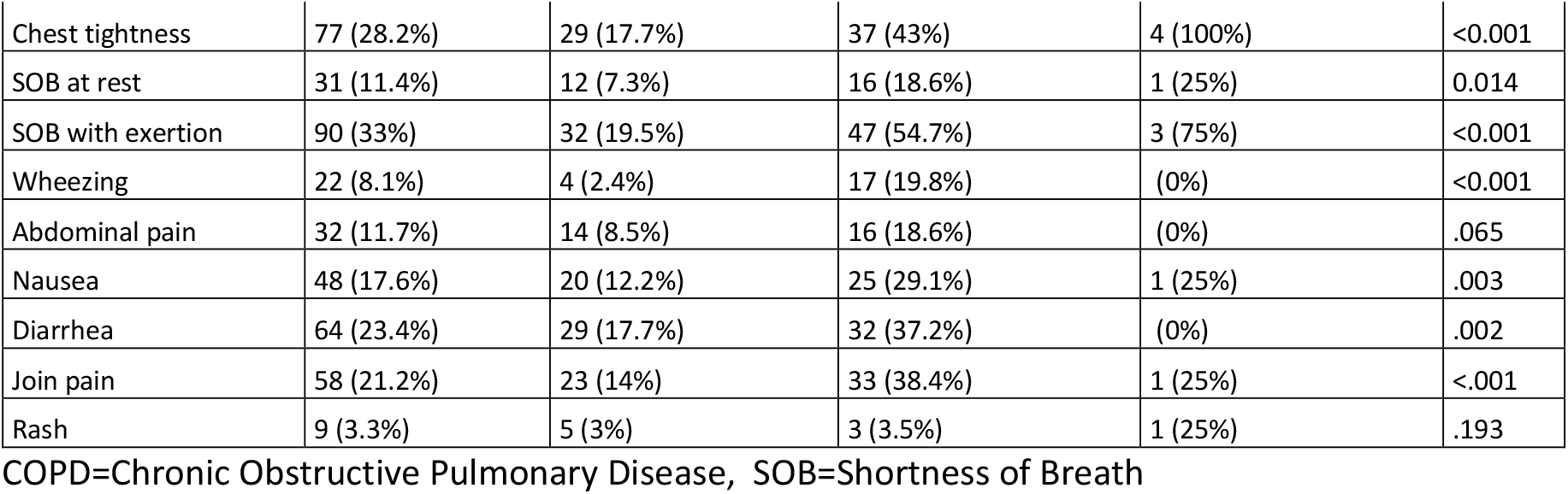
Demographics, comorbidities and symptoms at initial visit.

### Symptoms at initial visit

The symptoms reported by the cohort at the intake visit included 64% cough, 53% loss of smell or taste, 50% sinus congestion, 50% headache, 48% body aches, 36% chills, 33% shortness of breath with exertion, 28% chest tightness, 23% diarrhea, 22% dizziness, 22% sore throat, 22% joint pain, 18% fever, 11% shortness of breath, 8% wheezing, 3% rash (Table 1). The groups differed significantly except for rash, abdominal pain, and sinus congestion.

### Time course of individual symptoms

Figure 1 displays the heat map of symptoms over a 30 day follow up period for all 273 patients by percentage of patients with symptoms. Among all patients, the most prevalent symptom reported during 30 days of follow up was cough (62%), loss of smell or taste (54%), body aches (52%), headache (50%), and nasal congestion (47%). The most frequent remaining symptoms at 30 days were cough (7%), loss of smell or taste (5%), body aches (5%), nasal congestion (5%), shortness of breath with exertion (5%), and joint pain (5%). Fever was not a prominent symptom during 30 days of follow up, present in 27% of patients early in the course of illness.

**Figure 1:**
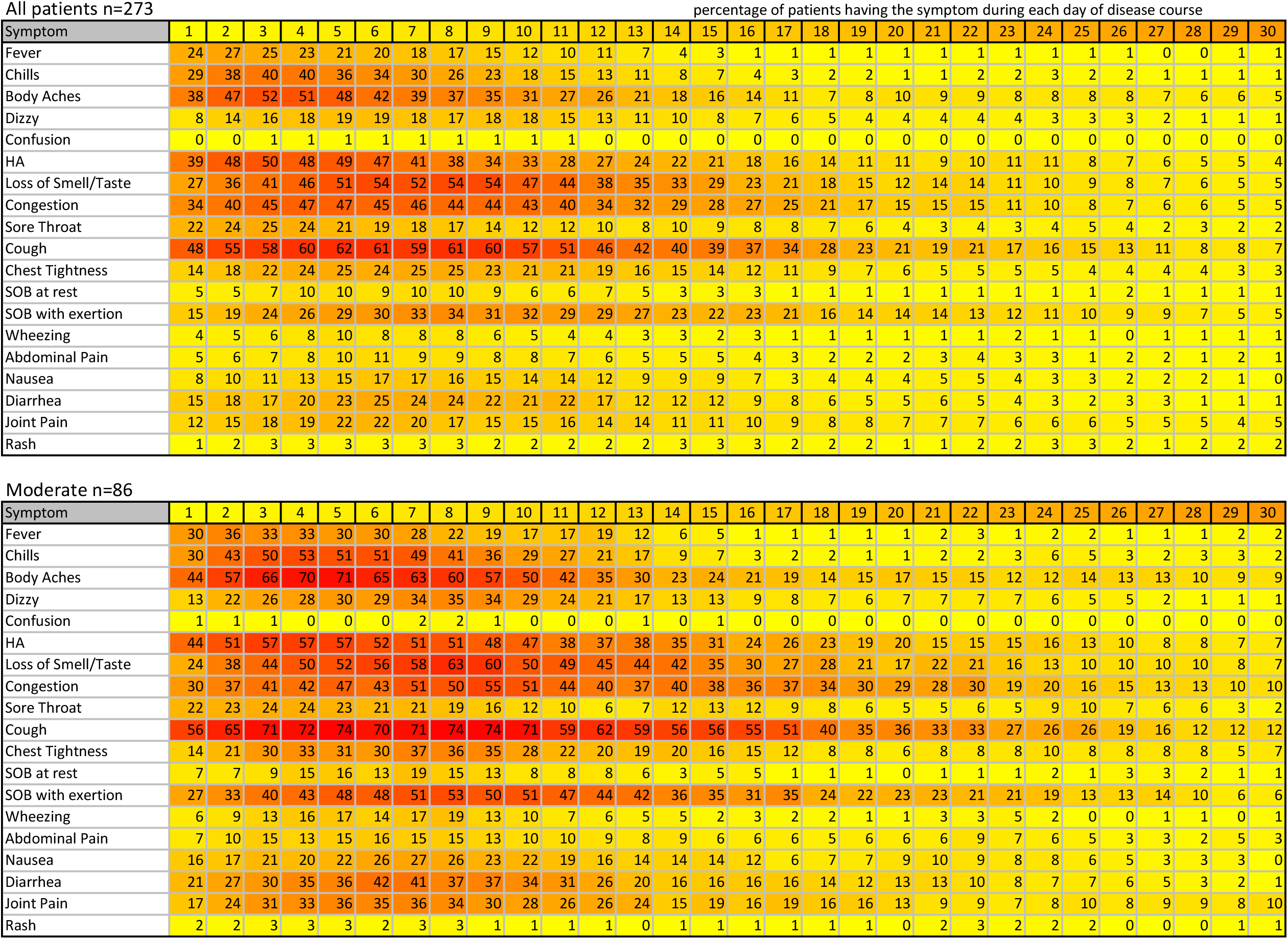
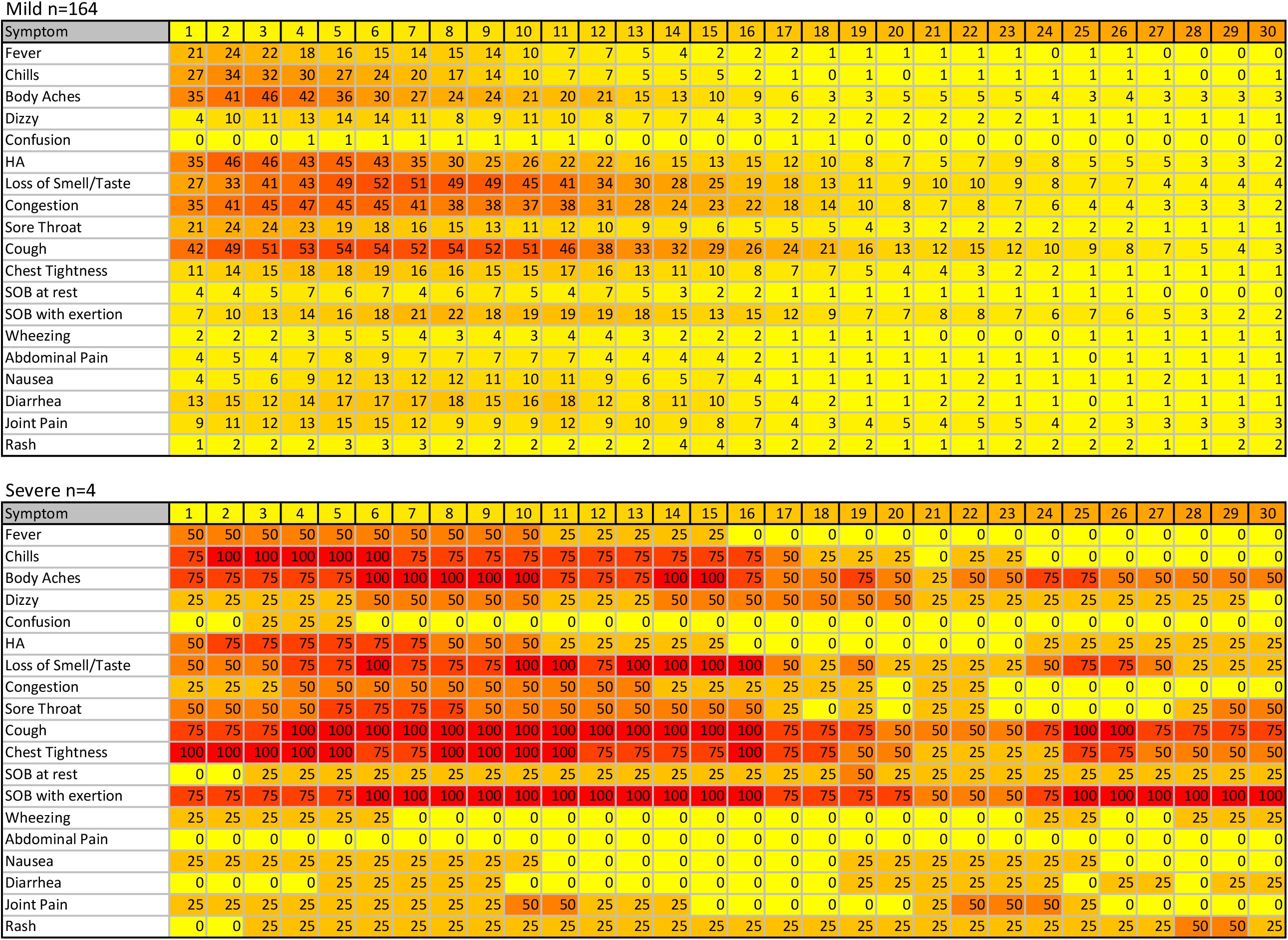
Time course of individual symptoms. Figure 1a: All patients, n=273 (% patients having symptom each day of covid-19 disease) Figure 1b: Mild provider assessed symptom severity n=164 Figure 1c: Moderate provider assessed symptom severity n=86 Figure 1d: Severe provider assessed symptom severity n=4

### Time course of symptoms by Provider-Assessed Symptom Severity

The heat map findings for Mild PASS was similar to the entire cohort, with loss of smell or taste in 52% over 30 days and 4% at 30 days and cough in 54% over 30 days and 3% at 30 days of follow up (Figure 1). At 30 days, 3% of patients described both body aches and joint pain. The heat map for Moderate PASS had higher rates of cough (74%), body aches (71%), loss of taste or smell (60%), headache (57%), nasal congestion (55%), shortness of breath with exertion (53%), chills (53%), fever (36%), diarrhea (42%), joint pain (36%) and dizziness (35%). At 30 days the most prominent symptom remaining was cough (12%), nasal congestion (10%), joint pain (10%), body aches (9%), loss of taste or smell (7%), headache (7%), and shortness of breath with exertion (6%). The heat map for the four patients with Severe PASS shows 75% of patients experienced chills, body aches, headache, loss of smell or taste, sore throat, cough, chest tightness, and shortness of breath with exertion during 30 days of follow up. At 30 days, 3 patients (75%) still had cough.

### Timing of symptom onset of by system

The day of symptom onset was earliest for upper respiratory symptoms, followed by systemic symptoms, with later onset of lower respiratory and gastrointestinal symptoms, when present (figure 2).

**Figure 2:**
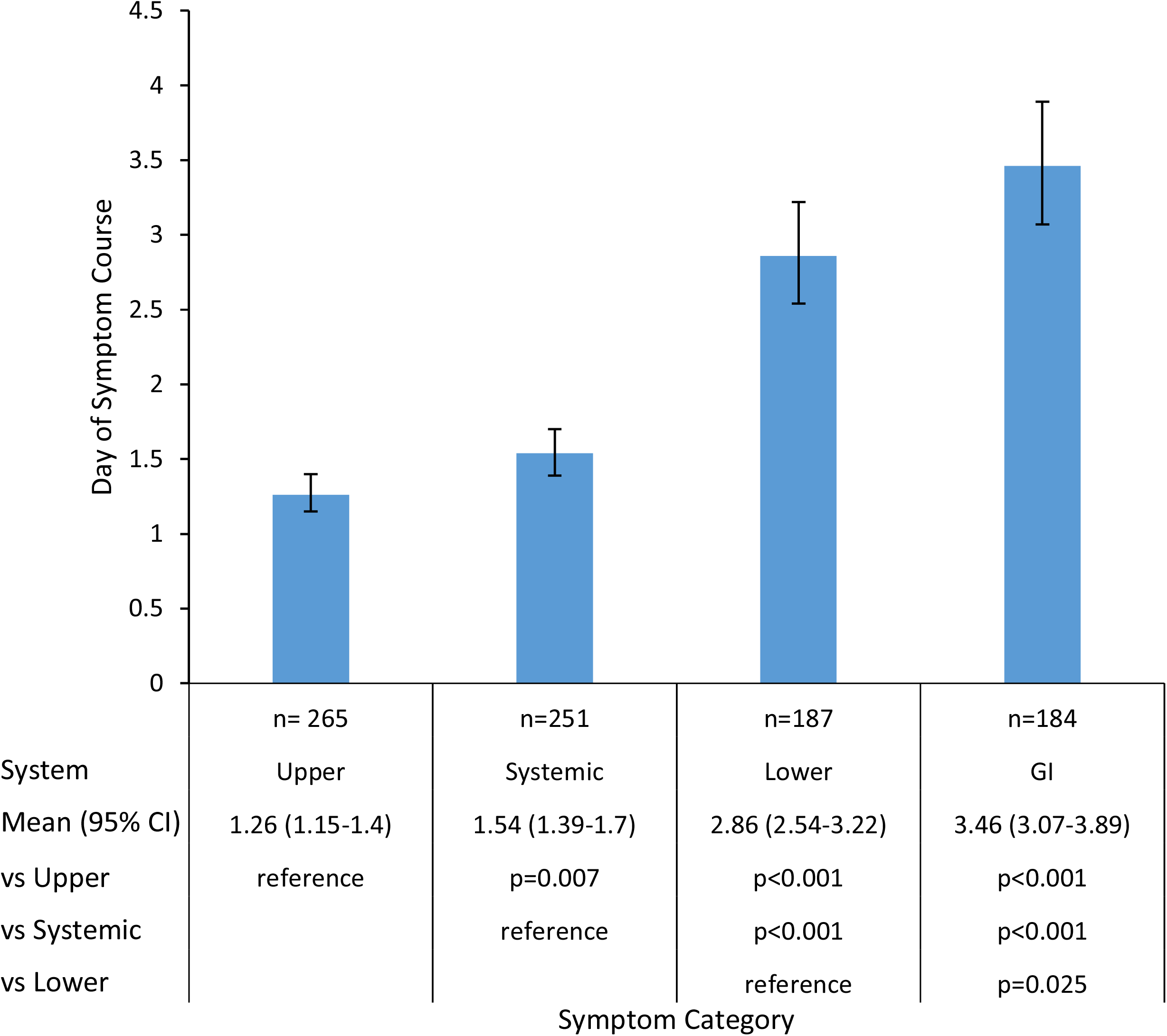
Day of Symptom Onset by System.

### Duration of each symptom

Figure 3 describes the mean days and 95% confidence interval for each symptom followed in the cohort over 30 days. Cough had the longest duration with 12.2 days (95% confidence interval 10.9 to 13.6, N=223). Loss of Smell or Taste had the second longest duration with 11.0 days (9.9 to 12.2, N=196).

**Figure 3:**
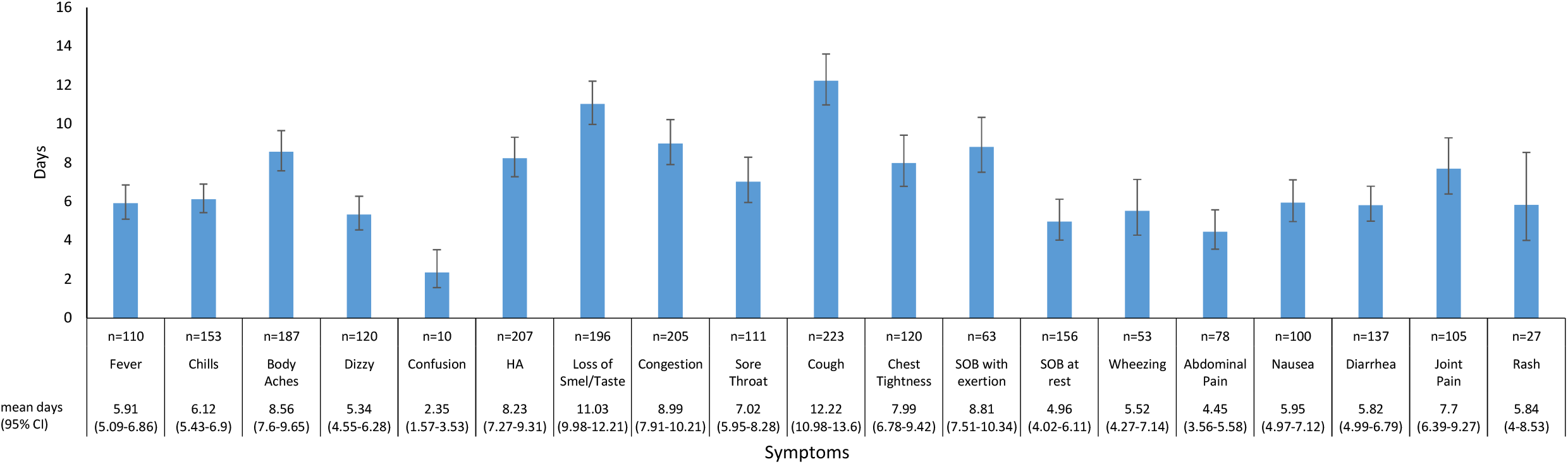
Duration of Symptoms.

### Symptom duration by Provider-Assessed Symptom Severity

PASS and patient-reported symptom severity were the only two predictors of disease duration that met prespecified statistical significance with p < 0.005. PASS was significantly correlated with more symptoms on ANOVA and, thus, was chosen for the model. No other predictors, including demographics and comorbidities, met significance for more than a single symptom and therefore multi-way ANOVA was not performed. Figure 4 shows the duration of symptoms for the overall group alongside the duration for each PASS group. Body aches, shortness of breath with exertion, headache, diarrhea and congestion had significant difference in durations between mild and more severe PASS groups. Chest tightness had significant difference in duration in moderate versus severe PASS. Body aches, shortness of breath with exertion, dizziness and chest tightness did not reach the level of statistical significant we set out in methods likely due to the small number of patients in the severe PASS group. Loss of smell or taste also did not reach the threshold set out in methods section but we kept it to compare with other publications.

**Figure 4:**
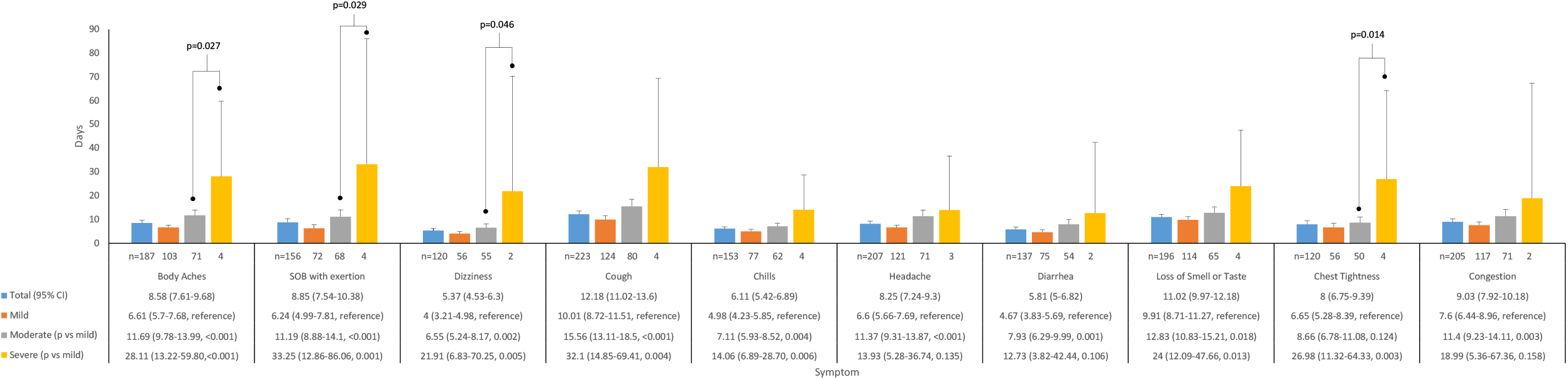
Symptom Duration by Provider Assessed Symptom Severity.

## DISCUSSION

### Principal findings

In this cohort of non-hospitalized patients with covid-19, disease course follows a pattern of progression that is illustrated with visual heatmaps of symptom frequency over time. The most common initial symptoms are systemic, upper respiratory and cough. Lower respiratory and gastrointestinal symptoms are less frequent and have a later onset in the disease course. The symptoms with the longest duration, when present, are cough, loss of smell or taste, sinus congestion, shortness of breath on exertion, body aches, and headache.

Patients were assigned a symptom severity (PASS) at the intake visit using a tool primarily aimed at determining hospitalization risk,[19] which was therefore based primarily on respiratory symptoms. Consistent with this, PASS is significantly associated with the presence of lower respiratory symptoms such as cough, chest tightness and shortness of breath with exertion. However, PASS was also associated with systemic symptoms (chills, body aches and dizziness) as well as greater incidence of symptoms from other organ systems (headache, loss of smell or taste, nausea, diarrhea). With clinical experience, it is possible that providers developed knowledge of other symptom categories and integrated these into the PASS. It is notable that sinus congestion, a common upper respiratory complaint, did not differ significantly between provider-rated severity groups.

An important observation in this study is that patient factors used to predict risk of severe disease (age, comorbidities) did not predict disease duration. Instead, we find that initial provider assessment of symptom severity is the best predictor of disease duration. PASS predicts duration for a number of symptoms, with significant associations for respiratory symptoms as well as non-respiratory symptoms (including body aches, dizziness, chills, headache, and diarrhea). This highlights a role for clinical providers in the assessment of acute covid-19 and providing expectant counseling that may be difficult to replace with automated patient monitoring systems using self-reported symptom severity.

### Comparison with other studies

Initial symptoms reported are similar to previous studies of mild covid-19 and non-hospitalized subsets.[12, 15] It differs from the overall reported literature, summarized in a recent systematic review of 148 studies.[14] Notably, fever was less common (n=49, 17.9%) compared with 78% in the systematic review, while other symptoms are more common, for example: headache (50.2% vs 13%), body aches (47.6% vs 17%), hyposmia (53.1% vs 25%). The higher frequency of multiple symptoms may be due to our systematic approach to symptom inquiry. Rate of fever may be underreported due to template wording “current fever” but also may be less frequent in this cohort as increased testing availability has expanded the symptom profile of “mild covid-19” patients eligible for screening.

We find that the course of illness and predictors of symptom duration have not been well described and this is an important contribution of this analysis. Narrative reviews have noted symptom progression similar to our report[21] and the visual course reported here in heatmap form illustrates the development of respiratory symptoms during and after the first week of illness among patients never requiring hospitalization for covid-19.

The persistence of symptoms identified in our study is also an important finding for clinical practice. Patients and providers may be reassured that gradual resolution is typical based on our findings. In our experience providing in-person care, many patients present for evaluation of non-resolving symptoms during subacute or convalescent illness.[22] Other reports have noted long duration of medical leave among persons with covid-19, for example first responders in New York (leave duration mean 25.3 days, SD=13.2).[23] We are able to differentiate the likelihood of prolonged symptoms in patients using mild and moderate PASS, which may aid in clinical counseling and anticipation of symptom recovery times. Given reports of delayed recovery of symptoms after hospitalization,[24] the differentiation by symptom severity in outpatients is plausible.

### Strengths and limitations of study

Our data on symptom course in outpatients is robust due to the structure of the VOMC, which was staffed to meet the anticipated “surge” of patients in March-May 2020 and therefore had skilled providers contacting patients and completing full note templates regularly through the course of acute illness. Missing clinical data were minimal (e.g. low risk patients contacted every 48 hours instead of 24 hours), allowing for standard approach to imputation.

The primary limitation of this study is that it represents a single-center cohort of patients screened during the early SARS-CoV-2 pandemic. Screening criteria favored the inclusion of working-age individuals in the cohort and our exclusion of hospitalized patients favors younger and healthier patients. We have limited numbers of patients with comorbidities and cannot therefore draw conclusions about the duration of symptoms related to specific conditions (e.g. chronic obstructive pulmonary disease).

Another limitation of the structured VOMC cohort data is the time to intake visit. Usual care requires a positive SARS-CoV-2 test prior to enrollment, and delays in testing could attenuate recall of initial symptoms. We therefore limited the study to patients within 10 days of symptom onset and used chart review to verify symptoms reported in the screening process. Discharge timing in the VOMC was a limitation for our follow-up data: the VOMC discharge criteria mirrored the CDC terminology of symptom “improvement,” but not resolution. We find in other work (unpublished data) that minor residual symptoms are common after VOMC discharge (reported in 55 of 158, 34.8%, of patients contacted a mean of 37.9 days after discharge) and that few (n=7, 4.4%) have symptoms requiring medical follow-up (e.g. by a primary care physician or specialist).[25] These residual symptoms are not captured in the heatmap data after their final VOMC call.

## CONCLUSION

Overall, we find that the symptom course of outpatients with covid-19 follows a pattern described in early observations with a typical illness course progressing from early symptoms (systemic, upper respiratory, and cough) to lower respiratory and gastrointestinal symptoms. We confirm that symptoms of altered smell or taste and headache are common in outpatients. Prolonged symptoms are common and the severity of symptoms in the acute phase of illness is the most significant predictor of disease duration.

## Data Availability

Deidentified data are available for sharing upon reasonable request.

https://dataverse.unc.edu/dataverse/VOMCTierStudy

## MANUSCRIPT INFORMATION

### Ethics

The study was approved by the Emory University Institutional Review Board (STUDY00000766), which granted both a waiver of informed consent and a waiver of the Health Information Portability and Privacy Act as the study posed no more than minimal risk.

## Acknowledgments

We would like to acknowledge Dr. David Roberts, MD for the design of the structured intake assessment note and nurse follow-up notes. We would also like to acknowledge the members of the Virtual Outpatient Management Clinic including faculty, staff and administrative members of the Paul W. Seavey Comprehensive Internal Medicine Clinic and Emory at Rockbridge Primary Care clinic as well as the physicians, nurses, and advanced practice providers who volunteered from other sites.

## Data Sharing Statement

Deidentified data are available for sharing upon reasonable request.

## Author Contributions

The corresponding author attests that all listed authors meet authorship criteria and that no others meeting the criteria have been omitted. David Tong had full access to all the data in the study and take responsibility for the integrity of the data and the accuracy of the data analysis.

*Concept and design:* All authors

*Acquisition, analysis, or interpretation of data:* All authors

*Drafting of the manuscript:* JO, BT, DT

*Critical revision of the manuscript for important intellectual content:* All authors

*Statistical analysis:* DT

*Obtained funding:* N/A

*Administrative, technical, or material support:* N/A

*Supervision:* N/A

## Conflict of Interest Disclosures

All authors have completed the Unified Competing Interest form (available on request from the corresponding author) and declare: no support from any organisation for the submitted work; no financial relationships with any organisations that might have an interest in the submitted work in the previous three years, no other relationships or activities that could appear to have influenced the submitted work. Dr. G. O’Keefe served on an advisory board of Eyepoint Pharmaceuticals in 2019. It is unrelated to the current work.

## Funding/Support

N/A

## Role of Funder/Sponsor

N/A

## Meeting Presentations

Emory University Department of Medicine Research Day (2020)

## Notes

### Competing Interest Statement

Dr. G. OKeefe served on an advisory board of Eyepoint Pharmaceuticals in 2019. It is unrelated to the current work.

### Funding Statement

No external funding

### Summary of Updates

Additional analyses of (1) symptom duration by initial severity, (2) symptom onset timing

## REFERENCES

1. Argenziano MG, Bruce SL, Slater CL, Tiao JR, Baldwin MR, Barr RG, et al. Characterization and clinical course of 1000 patients with coronavirus disease 2019 in New York: retrospective case series. BMJ. 2020 May 29;369:m1996.

2. Docherty AB, Harrison EM, Green CA, Hardwick HE, Pius R, Norman L, et al. Features of 20?133 UK patients in hospital with covid-19 using the ISARIC WHO Clinical Characterisation Protocol: prospective observational cohort study. BMJ. 2020 May 22;369:m1985.

3. Huang C, Wang Y, Li X, Ren L, Zhao J, Hu Y, et al. Clinical features of patients infected with 2019 novel coronavirus in Wuhan, China. Lancet. 2020 Feb 15;395(10223):497– 506.

4. Wang D, Hu B, Hu C, Zhu F, Liu X, Zhang J, et al. Clinical Characteristics of 138 Hospitalized Patients With 2019 Novel Coronavirus–Infected Pneumonia in Wuhan, China. JAMA. 2020 Mar 17;323(11):1061–9.

5. Zhou F, Yu T, Du R, Fan G, Liu Y, Liu Z, et al. Clinical course and risk factors for mortality of adult inpatients with COVID-19 in Wuhan, China: a retrospective cohort study. Lancet. 2020 Mar 28;395(10229):1054–62.

6. Stokes EK, Zambrano LD, Anderson KN, et al. Coronavirus Disease 2019 Case Surveillance — United States, January 22–May 30, 2020. MMWR Morb Mortal Wkly Rep 2020;69:759–765. http://dx.doi.org/10.15585/mmwr.mm6924e2

7. Centers for Disease Control and Prevention. Interim Clinical Guidance for Management of Patients with Confirmed Coronavirus Disease (COVID-19). Updated March 7, 2020. Available: https://www.cdc.gov/coronavirus/2019-ncov/hcp/clinical-guidance-management-patients.html. Accessed March 18, 2020.

8. World Health Organization. Home care for patients with COVID-19 presenting with mild symptoms and management of their contacts. Updated March 17, 2020. Available: https://www.who.int/publications-detail/home-care-for-patients-with-suspected-novel-coronavirus-(ncov)-infection-presenting-with-mild-symptoms-and-management-of-contacts. Accessed March 18, 2020.

9. Matthew F Pullen, Caleb Skipper, Kathy H Hullsiek, Ananta S Bangdiwala, Katelyn A Pastick, Elizabeth C Okafor, Sarah M Lofgren, Radha Rajasingham, Nicole Engen, Alison Galdys, Darlisha Williams, Mahsa Abassi, David R Boulware, Symptoms of COVID-19 Outpatients in the United States, Open Forum Infectious Diseases,, ofaa271, https://doi.org/10.1093/ofid/ofaa271

10. Lapostolle F, Schneider E, Vianu I, Dollet G, Roche B, Berdah J, et al. Clinical features of 1487 COVID-19 patients with outpatient management in the Greater Paris: the COVID-call study. Intern Emerg Med. 2020 May 30;28395(10229):1054.

11. Kluytmans-van den Bergh Mfq, Buiting AGM, Pas SD, Bentvelsen RG, van den Bijllaardt W, van Oudheusden AJG, et al. Prevalence and Clinical Presentation of Health Care Workers With Symptoms of Coronavirus Disease 2019 in 2 Dutch Hospitals During an Early Phase of the Pandemic. JAMA Netw Open. 2020 May 1;3(5):e209673.

12. Kim GU, Kim MJ, Ra SH, Lee J, Bae S, Jung J, et al. Clinical characteristics of asymptomatic and symptomatic patients with mild COVID-19. Clinical Microbiology and Infection. Elsevier Ltd; 2020 Jul 1;26(7):948.e1–948.e3.

13. Killerby ME, Link-Gelles R, Haight SC, Schrodt CA, England L, Gomes DJ, et al. Characteristics Associated with Hospitalization Among Patients with COVID-19 - Metropolitan Atlanta, Georgia, March-April 2020. MMWR Morb Mortal Wkly Rep. 2020 Jun 26;69(25):790–4.

14. Grant MC, Geoghegan L, Arbyn M, Mohammed Z, McGuinness L, Clarke EL, et al. The prevalence of symptoms in 24,410 adults infected by the novel coronavirus (SARS-CoV-2; COVID-19): A systematic review and meta-analysis of 148 studies from 9 countries. Hirst JA, editor. PLoS ONE. 2020 Jun 23;15(6):e0234765–19.

15. Burke RM, Killerby ME, Newton S, Ashworth CE, Berns AL, Brennan S, et al. Symptom Profiles of a Convenience Sample of Patients with COVID-19 - United States, January-April 2020. MMWR Morb Mortal Wkly Rep. 2020 Jul 17;69(28):904–8.

16. Tabata S, Imai K, Kawano S, Ikeda M, Kodama T, Miyoshi K, et al. Clinical characteristics of COVID-19 in 104 people with SARS-CoV-2 infection on the Diamond Princess cruise ship: a retrospective analysis. Lancet Infect Dis. 2020 Jun 12.

17. Tenforde MW, Kim SS, Lindsell CJ, et al. Symptom Duration and Risk Factors for Delayed Return to Usual Health Among Outpatients with COVID-19 in a Multistate Health Care Systems Network — United States, March–June 2020. MMWR Morb Mortal Wkly Rep. ePub: 24 July 2020. DOI: http://dx.doi.org/10.15585/mmwr.mm6930e1

18. O’Keefe JB, Cellai M. Characterization of prolonged COVID-19 symptoms and patient comorbidities in an outpatient telemedicine cohort. medRxiv. 2020 Jan 1;:2020.07.05.20146886.

19. O’Keefe JB, Tong EJ, Taylor TH, Datoo O ‘ Keefe GD, Tong DC. Initial Experience in Predicting the Risk of Hospitalization of 496 Outpatients with COVID-19 Using a Telemedicine Risk Assessment Tool. medRxiv. 2020 Jan 1;:2020.07.21.20159384.

20. Williamson EJ, Walker AJ, Bhaskaran K, Bacon S, Bates C, Morton CE, et al. OpenSAFELY: factors associated with COVID-19 death in 17 million patients. Nature. 2020 Jul 8.

21. Cohen PA, Hall LE, John JN, Rapoport AB. The Early Natural History of SARS-CoV-2 Infection: Clinical Observations From an Urban, Ambulatory COVID-19 Clinic. Mayo Clinic Proceedings. 2020 Jun;95(6):1124–6.

22. Ramakrishnan A, Zreloff J, Moore M, Bergquist SH, Cellai M, Higdon J, O’Keefe JB, Roberts DL, Wu HM. Persistence of Respiratory and Non-Respiratory Symptoms among COVID-19 Patients Seeking Care at an Ambulatory COVID-19 Center. ID Week 2020. Virtual. October, 2020 (oral presentation).

23. Prezant DJ, Zeig-Owens R, Schwartz T, Liu Y, Hurwitz K, Beecher S, et al. Medical Leave Associated With COVID-19 Among Emergency Medical System Responders and Firefighters in New York City. JAMA Netw Open. 2020 Jul 24;3(7):e2016094–4.

24. Carfí A, Bernabei R, Landi F, Gemelli Against COVID-19 Post-Acute Care Study Group. Persistent Symptoms in Patients After Acute COVID-19. JAMA. 2020 Jul 9.

25. O’Keefe JB, Datoo O’Keefe GA, Mufarreh A. Short Paper: Risk Factors for Long-Term Persistent Symptoms in COVID-19 in an Outpatient Cohort. 2020. (submission)

